# Rehabilitation strategies for lateral ankle sprain do not reflect established mechanisms of re-injury: A systematic review

**DOI:** 10.1101/2022.09.10.22279799

**Authors:** Jente Wagemans, Chris Bleakley, Jan Taeymans, Kevin Kuppens, Alexander Philipp Schurz, Heiner Baur, Dirk Vissers

## Abstract

**Research questions:**

1. What is the primary impairment addressed by each exercise included in exercise-based rehabilitation programs for patients who suffered an acute ankle sprain?
2. Do prescribed exercises incorporate complex tasks associated with common ankle sprain injury mechanisms?

**Methods:** We searched six electronic databases (CINAHL, Web of Science, SPORTDiscus, Cochrane Register of Controlled Trials, PEDro, Google Scholar) for randomized controlled trials including patients with acute ankle sprains, managed through exercise-based rehabilitation. Exercises were analysed based on: the primary impairment(s) addressed (muscle strength, mobility, neuromuscular training, performance); direction of movement (uni-vs multiplanar); base of support (single vs double limb); weightbearing status (open vs closed chain); and use of a flight phase. (PROSPERO: CRD42020210858)

**Results:** We included fourteen randomized controlled trials comprising 177 exercises. Neuromuscular function was addressed in 44% of exercises, followed by performance tasks (23%), and muscle strengthening (20%). Exercises were limited to movements across the sagittal plane (48%), with 31% incorporating multiplanar movements. Weight bearing exercises were almost divided equally between single-limb (59/122) and double leg stance exercises (61/122). Eighteen percent of all exercises (34/177) incorporated a flight phase.

**Conclusions:** Rehabilitation after LAS largely comprises simple exercises in the sagittal plane that do not reflect established mechanisms of re-injury. Future interventions can be enhanced by incorporating more open chain joint position sense training, multiplanar single limb challenges, and jumping and landing exercises.

## Introduction

Lateral ankle sprains (LAS) are one of the most frequently occurring injuries among athletes.^1-3^ There is a high risk of LAS in volleyball, basketball and football with prevalence ranging from 26-41%.^3-5^ Athletes with a history of LAS have greater risk of future injury,^6^ with recurrence rates of 28-61% recorded in (Association) football^7^, American football, basketball, and volleyball.^3^ Around 40% of patients with a history of LAS have additional long-term problems, such as persisting pain, giving-way episodes, and perceived instability, termed chronic ankle instability (CAI).^8-10^

A 2022 systematic review with meta-analysis concluded that exercise-based rehabilitation reduces the risk of recurrent ankle sprain, compared to usual care.^11^ The authors also reported some inconsistencies in exercise content and dosage prescribed for LAS rehabilitation, and an optimal program remains unclear.^11, 12^ Researchers often simplify the exercise content within an experimental study to facilitate replication and increase participant adherence. However, if exercises are too basic, they lack context and specificity, particularly for athletic populations. Recent scoping reviews conclude that most rehabilitation exercises prescribed for common musculoskeletal injuries at the hip (Femero acetabulair Impingement)^13^ and knee (Anterior cruciale ligmant, Patellofemoral Pain Syndrome)^14, 15^, are too simplistic and lack complexity, specificity and progression.

A key clinical goal after an index LAS is to prevent recurrence.^16^ The ROAST consensus highlights the importance of addressing mechanical and sensorimotor insufficiencies after LAS^17^; which often present clinically as reduced mobility, strength, and postural control. Understanding the etiology, mechanisms and inciting events that lead to recurrent LAS are also crucial to effectively design rehabilitation exercises.^17^ Injury surveillance data suggest that most non-contact LAS occur during multidirectional, reactive phases of play, or during high-speed jumping and landing.^3^ Biomechanical studies show that recurrent LAS often involve excessive inversion and internal rotation at initial contact, and a delay in peroneus muscle activation.^18-21^ To date, no research has comprehensively analysed the nature of individual exercises that comprise popular LAS rehabilitation programs.

In a recent systematic review with meta-analysis, we reported about training volume after a comprehensive analysis of the included exercise programs. However, all extracted data regarding the included exercises was too extensive to fall withing the scope of that systematic review. Hence, we decided to perform a secondary exploratory literature review. No new RCT’s have emerged since the previous systematic review with meta-analysis was published. This exploratory literature review audited the content of exercise-based rehabilitation programs used in randomized controlled trials (RCT’s), with following objectives:

- Determine the primary impairment addressed by each exercise.
- Establish if prescribed exercises incorporate complex tasks associated with common LAS injury mechanisms (multiplanar movements, single limb stance, a flight phase) (phase of gait when both feet are off the ground at the same time).

## Methods

We performed a systematic literature search according to the Preferred Reporting Items for Systematic Reviews and Meta-analysis (PRISMA). The current literature review is a secondary analysis of the primary systematic review with meta-analysis.^11^ The protocol of the systematic review was a priori registered with the International Prospective Registration Register of Systematic Reviews (PROSPERO; CRD42020210858). Our 2021 systematic review and meta-analysis previously assessed the study quality and risk of bias of the included studies, and the therapeutic quality of the exercise programs.^11^

Databases PubMed, CINAHL, Web of Science, SPORTDiscus, Cochrane Central Register of Controlled Trials, Physiotherapy Evidence Database (PEDro) and Google Scholar were systematically searched to obtain relevant articles from inception throughout June 2022. Additionally, reference lists of screened articles were reviewed to find more relevant articles. Randomized controlled trials (RCT’s) which included acute ankle sprain patients who received an exercise-based rehabilitation intervention were eligible for inclusion. A detailed description of the search strategy, eligibility criteria and screening are published elsewhere.^11^

All individual rehabilitation exercises used across each of the included studies were extracted and analysed independently by two reviewers (JW & CB). Analysis was based on the author’s description of the exercise in the article text, figures and/or corresponding appendices or supplementary data. When illustrations or written details were not provided, or in the event of disagreement about allocation in the predetermined categories, a meeting was held to reach consensus between reviewers. Exercises were categorised to determine 1) the primary impairment addressed by each exercise, and 2) the extent to which prescribed exercises incorporated complex tasks associated with common LAS injury mechanisms. Exercises incorporating components of multiplanar movement, single limb weight-bearing, and a flight phase, were most reflective of injury etiology, and therefore presented the highest level of challenge to the patient. A priori definitions, developed from recent scoping reviews by Dischiavi et al.^13-15^ were used to categorize each exercise element.

### 1. Clinical impairments

Each exercise was categorised according to the clinical impairment that they primarily addressed. We acknowledge that exercises can target multiple clinical outcomes, but our objective was to ascertain the primary intent of the prescribing author in their exercise selection. The following clinical outcomes were considered: range of motion, strength, postural control and performance tasks. If applicable, subcategories were used to describe the direction of movement (Plantar flexion, Dorsi flexion, Inversion, Eversion).

### 2. Pathomechanics

Each exercise was analysed to determine (a) primary plane of movement, (b) weightbearing status, and (c) the presence of absence of a flight phase.

#### 2.a. Planes of movement

Initially, exercises were categorised as uniplanar (exercise occurred in uniquely in one plane of motion) or multiplanar (exercise occurred in two or three of the cardinal planes of motion). Uniplanar exercises were then categorised into sagittal, frontal, or transverse planes. Exercises such as heel raises, and toe raises are an example of isolated movements considered to occur primarily in the sagittal plane. A side jump is an example of a functional exercise considered to occur primarily in the frontal plane. Exercises such as inversion and eversion muscle strength training are an example of isolated movements considered to occur primarily in the transverse plane. If an exercise was identified as multiplanar, the multiplanar box was checked, and then the 2 or 3 planes were also identified in the analysis.

#### 2.b. Weightbearing Status

Single limb stance exercises involve bearing full weight on one extremity and contacting the ground. We considered an exercise with a sequential movement, whereby one foot moved in a step-by-step fashion, as having both phases of stance. Similarly, variations of lunges were classified as bilateral weight bearing exercises, because both feet were on the ground during the intentional phase of the exercise. An example of a single limb stance exercise is a single leg heel raise or single leg squat. In a non-weightbearing exercise, neither lower extremity is functionally upright (hip over knee over foot) with the foot/feet on the ground. Quadruped exercises, bridging, open chain ankle strengthening, were not considered weightbearing since the method and position of delivery was not reflective of the upright position identified in the injury mechanism.

#### 2.c. Flight Phase

For a flight phase to be present, the exercise must include a phase where both lower extremities are simultaneously off the ground during the exercise. This would include any running, jumping, or hopping variations.

## Results

The systematic literature search yielded 14 RCT’s after duplication and screening for eligibility. Figure 1 shows the Prisma Flow diagram.

**Figure 1.**
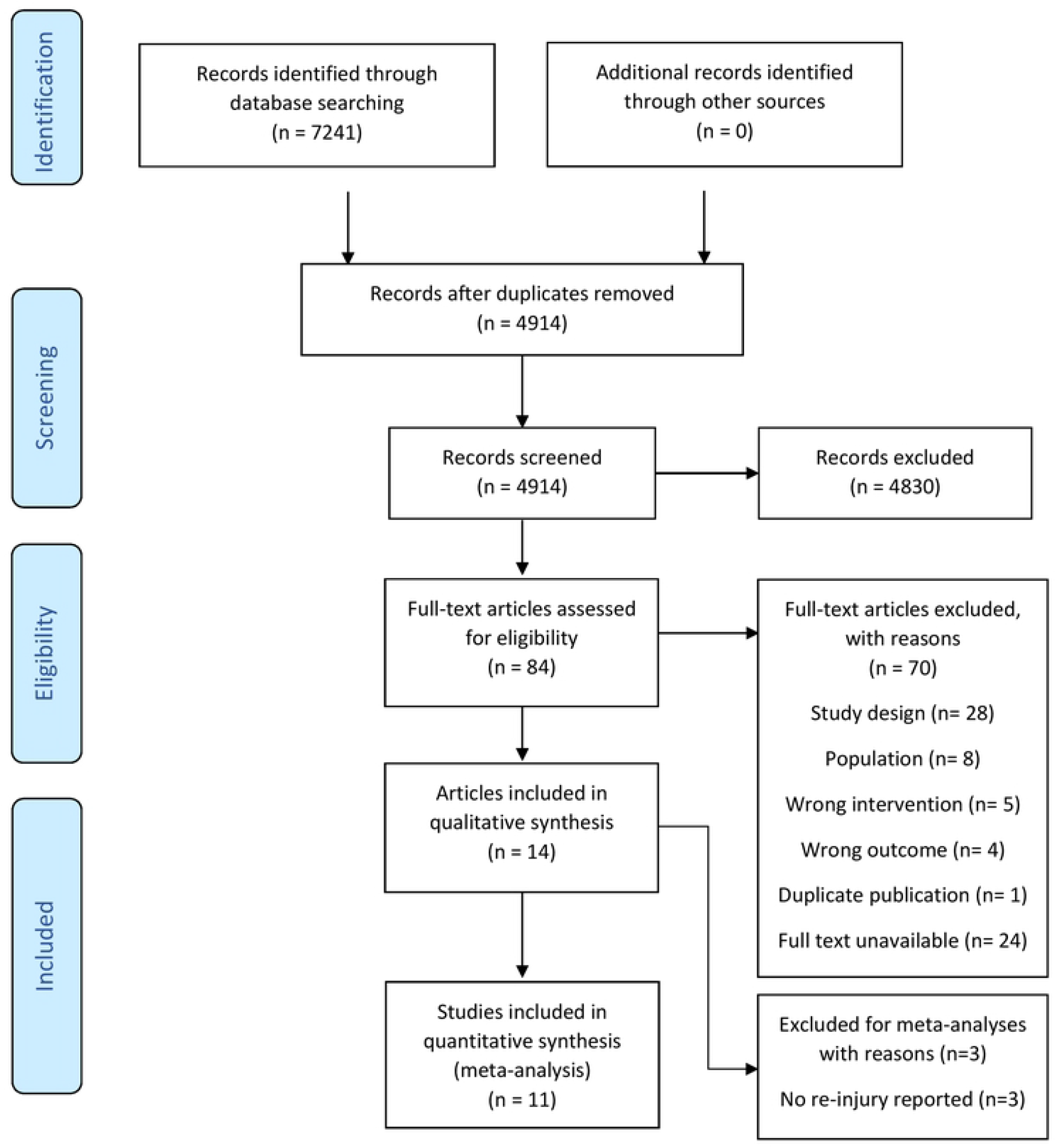
Prisma flow diagram.

**Figure 1a:**
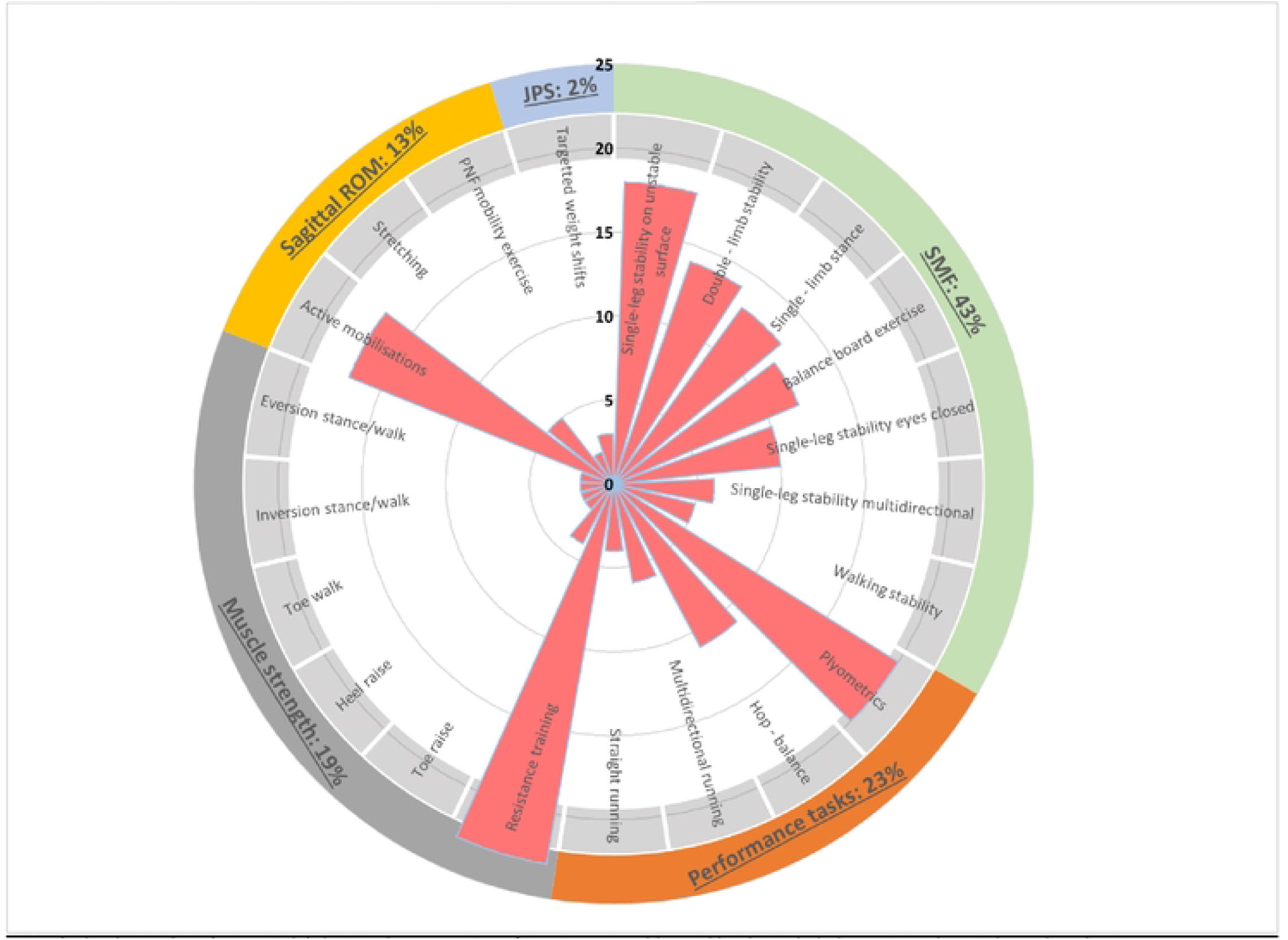
Clinical impairments with corresponding exercises (N=177) **Legend:** The donut chart (outer circle) depicts the proportion of impairments addressed by the included exercises; the wind rose bar chart (pink bars initiating from the centre) shows the different exercises that reflect each impairment and their number of occurrence. **Abbreviations**: SMF= Sensory motor function; ROM = Range of Motion; JPS= Joint position sense

**Figure 2:**
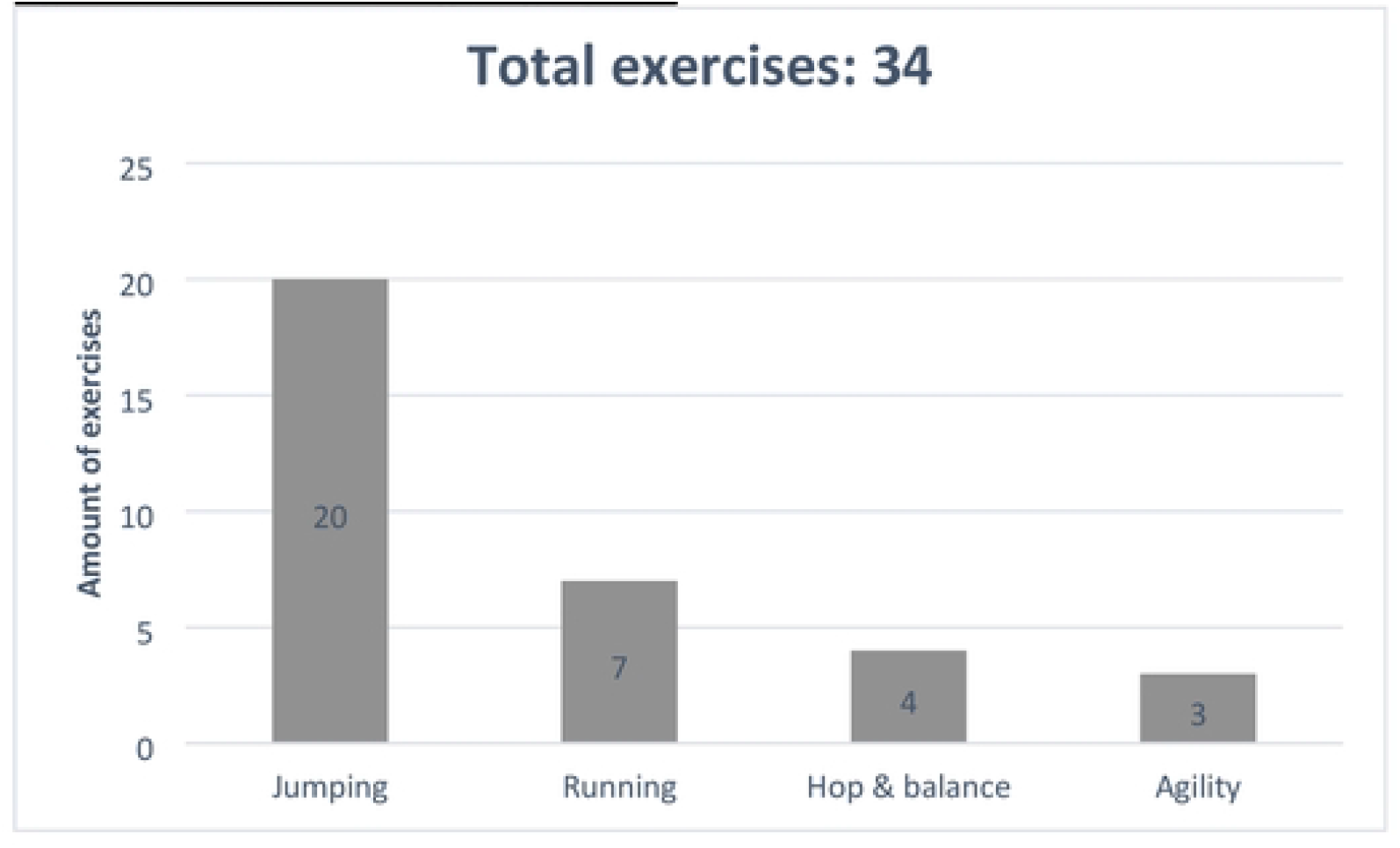
Clinical impairments with corresponding exercises (N=177) **Legend:** The donut chart (outer circle) depicts the proportion of impairments addressed by the included exercises; the wind rose bar chart (pink bars initiating from the centre) shows the different exercises that reflect each impairment and their number of occurrence. **Abbreviations**: SMF= Sensory motor function; ROM = Range of Motion; JPS= Joint position sense

**Figure 3:** Exercises including a flight phase.

### Study characteristics

Table 1 summarises the study characteristics. We included 14 randomized controlled trials, and a total of 177 exercises were extracted from the articles. The number of exercises prescribed within individual RCTs varied from three to 41. Time since injury at recruitment ranged from the day of injury to 11 weeks. Grade II ankle sprain were mostly represented. Five studies^22-26^ failed to mention injury severity. Five studies^23, 26-29^ included only athletes, and the population of two other studies^30, 31^ mostly comprised athletes.

**Table 1:** Study characteristics.

| Study | Participants <sup>A</sup> | Exercises (n) | Time since injury at recruitment | Injury severity (n) | Athletic vs non athletic |
| --- | --- | --- | --- | --- | --- |
| **Bleakley<sup>30</sup>** | n= 101; 69-32 (68-32%)<br>26y, ± 8y | 8 | <7 days | Grade 1 (29)<br>Grade 2 (73) | 62 – 39 |
| **Brisson<sup>32</sup>** | n= 503; 223-280 (44-56%)<br>30y, ± 13y | 30 | <72 hours | Grade 1 (149)<br>Grade 2 (354) | 215 – 288 |
| **Holme<sup>27</sup>** | n= 71; 44-27 (62-38%)<br>26y, ± 4y | 7 | Day of injury | Grade 1 (21)<br>Grade 2 (38)<br>Grade 3 (12) | 71 – 0 |
| **Hultman<sup>22</sup>** | n= 65; 35-30 (54-46%)<br>35y, ± 14y | 23 | Day of injury | Not stated | Not stated |
| **Hupperets<sup>23</sup>** | n= 522 ; 274-248 (52-48%)<br>28 y, ± 12 y | 7 | <2 months | Not stated | 522 – 0 |
| **Janssen<sup>28\*</sup>** | n= 340; 157-183 (46-54%)<br>34 y, ± 13 y | 7 | <2 months | Grade 1 (68)<br>Grade 2/3 (272) | 384 – 0 |
| **Kachanathu<sup>33</sup>** | n= 40; 24-16 (60-40%)<br>21.8 y, ± 2.9 y | 8 | Not stated | Grade 2 LAS | Not stated |
| **Lazarou<sup>24, 25</sup>** | n=20 ; 6-14 (30-70%)<br>22 y, ± 3 y | 10 | ≥ 11 weeks | Not stated | 10 – 10 |
| **Mohammadi<sup>26\*</sup>** | n= 60; 60-0 (100-0%)<br>24.6 y, ± 2.63 y | 3 | Not stated | Not stated | 60 – 0 |
| **Punt<sup>31\*</sup>** | n=90 ; 51-39 (64-36%)<br>32.7 y, ± 11 y | 22 | 4 weeks | Grade 1 (55)<br>Grade 2 (35) | 72 – 18 |
| **Van Reijen<sup>29</sup>** | n= 220; 110-110 (50-50%)<br>37.9 y, ± 13.4 y | 7 | <2 months | Grade 1 (91)<br>Grade 2 (64)<br>Grade 3 (18) | 220 – 0 |
| **Van Rijn<sup>34</sup>** | n= 102; 59-43 (58-42%)<br>37 y, ± 11.9 y | 41 | <1 week | Grade 1 (43)<br>Grade 2 (42)<br>Grade 3 (4) | Not stated |
| **Wester<sup>35</sup>** | n= 48; 29-19 (60-40%)<br>25 y, ± 7 y | 4 | Day of injury | Primary LAS; grade 2 | Not stated |

<sup>A</sup> Participants: total amount; male-female (%); mean age, ± SD Abbreviations: n= amount; SD: Standard deviation

### Exercise analysis

#### 1. Clinical impairments

Figure 1 summarises the clinical impairments addressed. About 44% (78/177) of all exercises primarily addressed postural control. Forty-one exercises addressed performance tasks. These exercises generally comprised plyometrics, jumping and landing, and running. Muscle strength was addressed in 20% of all exercises, with the majority focused on isolated strength training with resistance bands. ROM was addressed in 13 plantar flexion exercises and 12 dorsiflexion exercises. Exercises specifically addressing joint position sense (JPS) were underrepresented (3/177).

#### 2.a. Planes of motion

Table 2 depicts details regarding planes of motion. Most exercises were of uniplanar nature, with the sagittal plane most often represented. Most multiplanar exercises were biplanar by nature, and all multiplanar exercises comprised the sagittal plane.

**Table 2:**
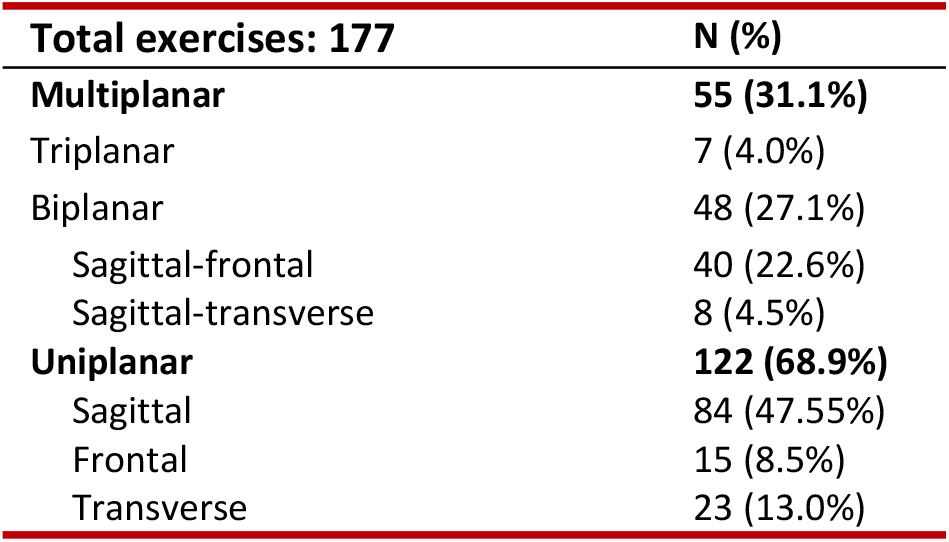
Planes of motion.

| Total exercises: 177 |  | N (%) |
| --- | --- | --- |
| **Multiplanar** |  | **55 (31.1%)** |
| Triplanar |  | 7 (4.0%) |
| Biplanar |  | 48 (27.1%) |
| Sagittal-frontal |  | 40 (22.6%) |
| Sagittal-transverse |  | 8 (4.5%) |
| **Uniplanar** |  | **122 (68.9%)** |
| Sagittal |  | 84 (47.55%) |
| Frontal |  | 15 (8.5%) |
| Transverse |  | 23 (13.0%) |

#### 2.b. Weightbearing Status

Exercises performed while standing were divided in single limb stance (59 exercises) and double leg stance exercises (61 exercises). The most common exercise performed standing involved closed kinetic chain strengthening (27 exercises). Twenty-seven exercises were closed kinetic chain strengthening exercises. Exercises that were not undertaken whilst bearing weight involved mostly exercises whilst seated, open kinetic chain exercises or exercises with a flight phase.

#### 2.c. Flight Phase

Only 34/177 exercises included a flight phase. Most were various types of jumping exercises (59%), such as high jumps, long jumps or skate jumps. Agility (9%) and hop & balance (12%) exercises were the least reported.

## Discussion

In this exploratory systematic review we audited the content of exercise-based rehabilitation programs in randomized controlled trials (RCT’s) that included patients with LAS. Previous reviews and guidelines^16, 36-38^ conclude exercise-based rehabilitation is effective after LAS, but because exercise content can vary, there is no consensus on an optimal rehabilitation program.^11, 12^ In this study, we reviewed individual rehabilitation exercises for LAS, categorizing them by the impairment addressed, and the extent to which each exercise reflects common mechanisms of re-injury. We included 14 RCT’s and categorized an aggregate of 177 exercises. A key finding was that most rehabilitation exercises prescribed within the literature are basic, consequently, some common impairments are not adequately addressed, and few exercises reflect established mechanisms of re-injury.

Ligament injuries such as LAS alter mechanoreceptor function, diminishing proprioception, balance, muscle strength and prolonging muscle reaction time.^39^ Our review found that almost half of exercises prescribed after LAS involve single leg balancing, which were often varied by standing on different surfaces and/or adding implicit motor challenges eg. catching a ball. This type of training integrates sensory information from visual, vestibular and proprioceptive fields,^40, 41^ and provides a safe medium for addressing neuromuscular deficits after LAS. Optimal rehabilitation should also involve a series of progressions, whereby exercises eventually recreate more vulnerable conditions and environments. A common inciting event for LAS is excessive ankle supination angle at initial foot contact.^3^ These errors are underpinned by aberrations in biomechanical position and (pre)activation of the muscles prior to ground contact; these can be addressed initially through explicit joint repositioning (JPS) tasks in the open kinetic chain. This approach has been used successfully during sensorimotor training at the shoulder joint^42, 43^ but surprisingly, we found that only 1.5% of exercises specifically involved open chain training at the ankle complex.

It was also surprising that just over 18% of all exercises included a flight phase. These consisted primarily of various jump exercises (20/34), such as single-leg jumps, forward jumps, stationary jumps and side jumps. Seven of 34 exercises that included a flight phase were running based, of which most involved straight line running, with agility exercises the least represented (3/34). These findings are also disproportionate to the (re)injury mechanism, as most non-contact LAS usually occur during jump landings, multidirectional running and sudden changes of direction.^18^ To adequately prepare the patient for these challenges, it is important that rehabilitation exercises move beyond closed chain postural control training, to include more reactive or “perturbation” training and stretch-shortening activities. Again, this should be done in a controlled and progressive fashion to ensure that neuromuscular adaptations occur without placing the patient at risk of re-injury.

A related limitation in the current literature, is that most exercises comprised uniplanar, sagittal dominant movements (47.6%). Common examples are dorsiflexion or plantarflexion exercises, calf stretching and heel or toe raises. Re-establishing sagittal plane mobility and strength is important for balance and functional performance tasks, such as running jumping and landing^44, 45^. However, exercises must also address impairments across other planes of movement. Non-contact LAS mechanisms are multidirectional, but only 31.1% of all exercises challenged more than one plane of motion, with the majority using sagittal-frontal plane combination. Non-contact LAS occur by an inversion-internal rotation, around the frontal and the transversal plane, with plantarflexion, around the sagittal plane.^9, 37^ Yet, only 4% of exercises challenged the three planes of motion. Restricting exercises to the sagittal plane also limits ligament recruitment and physiological adaptation.^46-48^ Incorporating multiplanar movements aligns more with the unique anatomy, morphology and biomechanical properties of the lateral ligaments. As connective tissues are mechano-responsive, inducing progressive and multidirectional loading through exercise also provide a medium to stimulate healing and alter tissue composition.^46^

Patients with an ankle sprain exhibit decreased eversion peak power,^49^ and reduced concentric and eccentric eversion peak torque are risk factors for the development of CAI.^50^ One-fifth of exercises targeted ankle muscle strength, but the majority were undertaken in the sagittal plane, with only 12/177 exercises (6.8%) primarily addressed eversion muscle strength. Peroneal reaction time can also be an influencing factor. Patients with a history of LAS exhibit a delayed peroneal reaction time, failing to protect the ankle from a sudden inversion perturbation.^51^ Exercises that addressed eversion muscle strength primarily involved concentric loading, using an elastic resistance band. Previous studies suggest that a 4-week elastic-resistance exercise-program failed to improve evertor muscle strength or peroneal latency, in patients with a history of LAS.^52^ The multiple degrees of freedom of movement, at the ankle complex can make it more difficult to isolate certain muscle groups, and optimal strength training may require additional patient supervision, to ensure optimal training technique.

Our main findings concur with similar scoping reviews undertaken in this field. Dischiavi et al.^15^ examined over 1000 exercises used for preventing ACL/knee injuries; they reported that most prescribed exercises did not address the task specific elements identified within ACL injury mechanisms,^15^ such as single leg landing, trunk hip dissociation, and multiplanar movements. Dischiavi and colleagues noted a reductionist approach to exercise prescription across other musculoskeletal conditions. Their scoping reviews of the patellofemoral pain (PFPS)^14^ and femoroacetabular impingement (FAI) literature,^13^ each concluded that the prescribed rehabilitation exercises were too simplistic, lacked progression, and often failed to reflect the etiology of the respective conditions. It seems there are consistent limitations in rehabilitation content across the current musculoskeletal literature; this could be addressed increasing the specificity, complexity and progression of the exercise content. The current approach limits the effect size of the intervention, particularly in athletes, where exercise progression is paramount for optimal rehabilitation.^53^ Exercise programs must be developed to incorporate more chaotic, sport-specific movements,^54^ which eventually replicate key features of injury mechanism.^53, 55^

## Limitations

Our key finding was that exercises prescribed after LAS do not incorporate complex tasks associated with common LAS injury mechanisms. Exercise elements were classified based on the impairment addressed, direction of movement, use of flight, and weight-bearing status. This classification could have been subjective; but to minimise this, we aligned our methods and definitions to three previous reviews.^13-15^ We must also acknowledge that our classification criteria is informed primarily by mechanical constructs relevant to the injury inciting event. This reductionist approach of discussing only biomechanical effects of rehabilitation exercises deviates from the original biopsychosocial model.^56^ Our criteria were not exhaustive and the etiology of re-injury involves a complex interaction of several internal and external risk factors^16, 57, 58^ These often include psychosocial impairments such as kinesiophobia, fear-avoidance, decreased quality of life, perceived instability and self-reported function.^9, 59, 60^ Current consensus guidelines such as ROAST and PAAS also highlight the importance of using patient-reported outcome measures in the rehabilitation process and return-to-sports decision making.^17, 61^ Future research should consider which psychosocial impairments are addressed by the prescribed exercises in rehabilitation programs.

## Conclusion

This review highlights that most rehabilitation exercises prescribed in current RCT’s are generic, simplistic, and do not fully reflect the pathomechanics of non-contact LAS. This could introduce a ceiling effect for LAS rehabilitation. Developing exercise interventions that better incorporate JPS training, multi-directional movements, flight phases and single limb landings, would present a more progressive task-specific approach to rehabilitation, cumulating in a greater reduction in re-injury risk. High quality prospective studies can determine the feasibility and clinical effectiveness of using more complex exercise-based training after acute LAS.

## Data Availability

People can send me an email if they want more data.

## Highlights

- Most rehabilitation exercises are generic and simplistic, across the sagittal plane.
- Key pathomechanics of a non-contact ankle sprain are not addressed by prescribed exercises in current RCT’s.

## Notes

### Competing Interest Statement

The authors have declared no competing interest.

### Funding Statement

The author(s) received no specific funding for this work.

## References

1. Gribble PA, Bleakley CM, Caulfield BM, et al. Evidence review for the 2016 International Ankle Consortium consensus statement on the prevalence, impact and long-term consequences of lateral ankle sprains. British journal of sports medicine. 2016;50(24):1496–505.

2. Fong DT, Hong Y, Chan LK, et al. A systematic review on ankle injury and ankle sprain in sports. Sports Med. 2007;37(1):73–94.

3. Herzog MM, Kerr ZY, Marshall SW, et al. Epidemiology of Ankle Sprains and Chronic Ankle Instability. J Athl Train. 2019;54(6):603–10.

4. Herzog MM, Mack CD, Dreyer NA, et al. Ankle Sprains in the National Basketball Association, 2013-2014 Through 2016-2017. The American journal of sports medicine. 2019;47(11):2651–8.

5. Verhagen EALM, Van der Beek AJ, Bouter LM, et al. A one season prospective cohort study of volleyball injuries. British journal of sports medicine. 2004;38(4):477–81.

6. Wikstrom EA, Cain MS, Chandran A, et al. Lateral ankle sprain increases subsequent ankle sprain risk: a systematic review. J Athl Train. 2020.

7. Attenborough AS, Hiller CE, Smith RM, et al. Chronic ankle instability in sporting populations. Sports Med. 2014;44(11):1545–56.

8. Doherty C, Bleakley C, Delahunt E, et al. Treatment and prevention of acute and recurrent ankle sprain: an overview of systematic reviews with meta-analysis. British journal of sports medicine. 2017;51(2):113–25.

9. Hertel J, Corbett RO. An Updated Model of Chronic Ankle Instability. J Athl Train. 2019;54(6):572–88.

10. McKeon PO, Donovan L. A Perceptual Framework for Conservative Treatment and Rehabilitation of Ankle Sprains: An Evidence-Based Paradigm Shift. J Athl Train. 2019;54(6):628–38.

11. Wagemans J, Bleakley C, Taeymans J, et al. Exercise-based rehabilitation reduces reinjury following acute lateral ankle sprain: A systematic review update with meta-analysis. PloS one. 2022;17(2):e0262023.

12. Bleakley CM, Taylor JB, Dischiavi SL, et al. Rehabilitation Exercises Reduce Reinjury Post Ankle Sprain, But the Content and Parameters of an Optimal Exercise Program Have Yet to Be Established: A Systematic Review and Meta-analysis. Arch Phys Med Rehabil. 2019;100(7):1367–75.

13. Wright AA, Tarara DT, Gisselman AS, et al. Do currently prescribed exercises reflect contributing pathomechanics associated with femoroacetabular impingement syndrome? A scoping review. Physical therapy in sport : official journal of the Association of Chartered Physiotherapists in Sports Medicine. 2021;47:127–33.

14. Dischiavi SL, Wright AA, Tarara DT, et al. Do exercises for patellofemoral pain reflect common injury mechanisms? A systematic review. Journal of science and medicine in sport. 2021;24(3):229–40.

15. Dischiavi SL, Wright AA, Heller RA, et al. Do ACL Injury Risk Reduction Exercises Reflect Common Injury Mechanisms? A Scoping Review of Injury Prevention Programs. Sports health. 2021:19417381211037966.

16. Vuurberg G, Hoorntje A, Wink LM, et al. Diagnosis, treatment and prevention of ankle sprains: update of an evidence-based clinical guideline. British journal of sports medicine. 2018;52(15):956.

17. Delahunt E, Bleakley CM, Bossard DS, et al. Clinical assessment of acute lateral ankle sprain injuries (ROAST): 2019 consensus statement and recommendations of the International Ankle Consortium. British journal of sports medicine. 2018;52(20):1304–10.

18. Li Y, Ko J, Zhang S, et al. Biomechanics of ankle giving way: A case report of accidental ankle giving way during the drop landing test. J Sport Health Sci. 2019;8(5):494–502.

19. Gehring D, Wissler S, Mornieux G, et al. How to sprain your ankle - a biomechanical case report of an inversion trauma. J Biomech. 2013;46(1):175–8.

20. Fong DT, Hong Y, Shima Y, et al. Biomechanics of supination ankle sprain: a case report of an accidental injury event in the laboratory. The American journal of sports medicine. 2009;37(4):822–7.

21. Lysdal FG, Wang Y, Delahunt E, et al. What have we learnt from quantitative case reports of acute lateral ankle sprains injuries and episodes of ‘giving-way’ of the ankle joint, and what shall we further investigate? Sport Biomech. 2022;21(4):359–79.

22. Hultman K, Fältström A, Öberg U. The effect of early physiotherapy after an acute ankle sprain. Advances in Physiotherapy. 2010;12(2):65–73.

23. Hupperets MD, Verhagen EA, van Mechelen W. Effect of unsupervised home based proprioceptive training on recurrences of ankle sprain: randomised controlled trial. Bmj. 2009;339:b2684.

24. Lazarou L, Kofotolis N, Pafis G, et al. Effects of two proprioceptive training programs on ankle range of motion, pain, functional and balance performance in individuals with ankle sprain. Journal of Back and Musculoskeletal Rehabilitation. 2018;31:437–46.

25. Lazarou L, Kofotolis N, Malliou P, et al. Effects of two proprioceptive training programs on joint position sense, strength, activation and recurrent injuries after ankle sprains. Isokinetics and Exercise Science. 2017;25:289–300.

26. Mohammadi F. Comparison of 3 preventive methods to reduce the recurrence of ankle inversion sprains in male soccer players. The American journal of sports medicine. 2007;35(6):922–6.

27. Holme E, Magnusson SP, Becher K, et al. The effect of supervised rehabilitation on strength, postural sway, position sense and re-injury risk after acute ankle ligament sprain. Scand J Med Sci Sports. 1999;9(2):104–9.

28. Janssen KW, van Mechelen W, Verhagen EA. Bracing superior to neuromuscular training for the prevention of self-reported recurrent ankle sprains: a three-arm randomised controlled trial. British journal of sports medicine. 2014;48(16):1235–9.

29. Van Reijen M, Vriend I, Zuidema V, et al. The “Strengthen your ankle” program to prevent recurrent injuries: A randomized controlled trial aimed at long-term effectiveness. Journal of science and medicine in sport. 2017;20(6):549–54.

30. Bleakley CM, O’Connor SR, Tully MA, et al. Effect of accelerated rehabilitation on function after ankle sprain: randomised controlled trial. Bmj. 2010;340:c1964.

31. Punt IM, Ziltener JL, Monnin D, et al. Wii Fit™ exercise therapy for the rehabilitation of ankle sprains: Its effect compared with physical therapy or no functional exercises at all. Scand J Med Sci Sports. 2016;26(7):816–23.

32. Brison RJ, Day AG, Pelland L, et al. Effect of early supervised physiotherapy on recovery from acute ankle sprain: randomised controlled trial. BMJ. 2016;355:i5650.

33. Kachanathu SJ, Hafez AR, Alenazi AM, et al. Wirksamkeit von Übungen in geschlossenen und offenen kinematischen Ketten zur Rehabilitation von Distorsionen der Knöchelregion. Physikalische Medizin, Rehabilitationsmedizin, Kurortmedizin. 2016;26(01):28–31.

34. van Rijn RM, van Os AG, Kleinrensink GJ, et al. Supervised exercises for adults with acute lateral ankle sprain: a randomised controlled trial. The British journal of general practice : the journal of the Royal College of General Practitioners. 2007;57(543):793–800.

35. Wester JU, Jespersen SM, Nielsen KD, et al. Wobble board training after partial sprains of the lateral ligaments of the ankle: a prospective randomized study. The Journal of orthopaedic and sports physical therapy. 1996;23(5):332–6.

36. Martin RL, Davenport TE, Fraser JJ, et al. Ankle Stability and Movement Coordination Impairments: Lateral Ankle Ligament Sprains Revision 2021. The Journal of orthopaedic and sports physical therapy. 2021;51(4):Cpg1–cpg80.

37. D’Hooghe P, Cruz F, Alkhelaifi K. Return to Play After a Lateral Ligament Ankle Sprain. Curr Rev Musculoskelet Med. 2020;13(3):281–8.

38. Caldemeyer LE, Brown SM, Mulcahey MK. Neuromuscular training for the prevention of ankle sprains in female athletes: a systematic review. Phys Sportsmed. 2020;48(4):363–9.

39. Zech A, Hubscher M, Vogt L, et al. Neuromuscular training for rehabilitation of sports injuries: a systematic review. Medicine and science in sports and exercise. 2009;41(10):1831–41.

40. Han J, Anson J, Waddington G, et al. The Role of Ankle Proprioception for Balance Control in relation to Sports Performance and Injury. BioMed research international. 2015;2015:842804.

41. Röijezon U, Clark NC, Treleaven J. Proprioception in musculoskeletal rehabilitation. Part 1: Basic science and principles of assessment and clinical interventions. Man Ther. 2015;20(3):368–77.

42. Rogol IM, Ernst G, Perrin DH. Open and closed kinetic chain exercises improve shoulder joint reposition sense equally in healthy subjects. J Athl Train. 1998;33(4):315–8.

43. E SE, Lin YL, J HK, et al. Joint position sense – There׳s an app for that. J Biomech. 2016;49(14):3529–33.

44. Drewes LK, McKeon PO, Kerrigan DC, et al. Dorsiflexion deficit during jogging with chronic ankle instability. Journal of science and medicine in sport. 2009;12(6):685–7.

45. Basnett CR, Hanish MJ, Wheeler TJ, et al. Ankle dorsiflexion range of motion influences dynamic balance in individuals with chronic ankle instability. Int J Sports Phys Ther. 2013;8(2):121–8.

46. Rein S, Hagert E, Hanisch U, et al. Immunohistochemical analysis of sensory nerve endings in ankle ligaments: a cadaver study. Cells, tissues, organs. 2013;197(1):64–76.

47. Noyes FR, DeLucas JL, Torvik PJ. Biomechanics of anterior cruciate ligament failure: an analysis of strain-rate sensitivity and mechanisms of failure in primates. The Journal of bone and joint surgery American volume. 1974;56(2):236–53.

48. Yu J, Wong DW, Zhang H, et al. The influence of high-heeled shoes on strain and tension force of the anterior talofibular ligament and plantar fascia during balanced standing and walking. Medical engineering & physics. 2016;38(10):1152–6.

49. Pourkazemi F, Hiller C, Raymond J, et al. Using Balance Tests to Discriminate Between Participants With a Recent Index Lateral Ankle Sprain and Healthy Control Participants: A Cross-Sectional Study. Journal of Athletic Training. 2016;51(3):213–22.

50. Thompson C, Schabrun S, Romero R, et al. Factors Contributing to Chronic Ankle Instability: A Systematic Review and Meta-Analysis of Systematic Reviews. Sports Med. 2018;48(1):189–205.

51. Hoch MC, Mckeon PO. Peroneal Reaction Time after Ankle Sprain: A Systematic Review and Meta-analysis. Medicine & Science in Sports & Exercise. 2014;46(3):546–56.

52. Han K, Ricard MD. Effects of 4 Weeks of Elastic-Resistance Training on Ankle-Evertor Strength and Latency. Journal of Sport Rehabilitation. 2011;20(2):157–73.

53. Blanchard S, Glasgow P. A theoretical model for exercise progressions as part of a complex rehabilitation programme design. British journal of sports medicine. 2019;53(3):139–40.

54. Taberner M, Allen T, Cohen DD. Progressing rehabilitation after injury: consider the ‘control-chaos continuum’. British journal of sports medicine. 2019;53(18):1132–6.

55. Otte FW, Millar SK, Klatt S. Skill Training Periodization in “Specialist” Sports Coaching-An Introduction of the “PoST” Framework for Skill Development. Frontiers in sports and active living. 2019;1:61.

56. Cormack B, Stilwell P, Coninx S, et al. The biopsychosocial model is lost in translation: from misrepresentation to an enactive modernization. Physiother Theory Pract. 2022:1–16.

57. Bahr R, Holme I. Risk factors for sports injuries--a methodological approach. British journal of sports medicine. 2003;37(5):384–92.

58. Verhagen E, van Dyk N, Clark N, et al. Do not throw the baby out with the bathwater; screening can identify meaningful risk factors for sports injuries. British journal of sports medicine. 2018;52(19):1223–4.

59. Houston MN, Van Lunen BL, Hoch MC. Health-related quality of life in individuals with chronic ankle instability. J Athl Train. 2014;49(6):758–63.

60. Houston MN, Hoch JM, Hoch MC. College Athletes With Ankle Sprain History Exhibit Greater Fear-Avoidance Beliefs. J Sport Rehabil. 2018;27(5):419–23.

61. Smith MD, Vicenzino B, Bahr R, et al. Return to sport decisions after an acute lateral ankle sprain injury: introducing the PAASS framework-an international multidisciplinary consensus. British journal of sports medicine. 2021.

